# SARS-CoV2 sero-survey among adults involved in health care and health research in Guinea-Bissau, West Africa

**DOI:** 10.1101/2021.03.06.21253046

**Authors:** Christine Stabell Benn, Alberto Salinha, Sabado Mendes, Carlos Cabral, Cesario Martins, Sebastian Nielsen, Ane Bærent Fisker, Frederik Schaltz-Buchholzer, Charlotte Sværke Jørgensen, Peter Aaby

## Abstract

**Background:** Many African countries have reported fewer COVID-19 cases than countries elsewhere. By the end of 2020, Guinea-Bissau, West Africa, had <2500 PCR-confirmed cases corresponding to 0.1% of the ∼1.8 million national population. We assessed the prevalence of SARS-CoV2 antibodies in urban Guinea-Bissau.

**Methods:** We measured IgG antibody in point-of-care rapid tests among 140 staff and associates at a biometric research field station in Bissau, the capital of Guinea-Bissau, during November 2020.

**Results:** Of 140 participants, 25 (18%) were IgG-positive. Among IgG-positives, 12 (48%) reported an episode of illness since the onset of the pandemic. Twenty-five (18%) participants had been PCR-tested between May and September; 7 (28%) were PCR-positive. Four of these 7 tested IgG-negative in the present study. Five participants reported a death in their house, corresponding to a crude annual death rate of 4.5/1000 people; no death was attributed to COVID-19.

**Conclusions:** In spite of low official number of COVID-19 cases, our serosurvey found a high prevalence of IgG-positivity. Most IgG-positives had not been ill. The official number of PCR-confirmed COVID-19 cases grossly underestimates the prevalence during the pandemic. The observed overall mortality rate is not higher than the official Guinean mortality rate of 9.6/1000 people.

## Background

In Guinea-Bissau, with a population of 1.9 million inhabitants, the first case of COVID-19 was registered on March 25, 2020, and was quickly followed by a lockdown that lasted several months. Per December 20, of 35,644 people tested by PCR, 2,447 (6.9%) tested positive for SARS-CoV2 (0.1% of the national population) with 45 deaths (1.8% of positive cases)(Figure[1]). We maintain a biomedical research field station in the capital Bissau. Most staff and associates had been working throughout the epidemic. We aimed to study the prevalence of SARS-CoV2 antibodies.

## Methods

The Bandim Health Project (www.bandim.org) employs ∼180 staff members and covers the suburbs Bandim, Belem, Mindara and Cuntum. We conducted a serosurvey among our local staff and associates: field assistants that conduct house visits to collect demographic and health information, office staff members, and staff placed at three health centers in the study area and the nearby national hospital.

From November 9-24, 2020, after informed oral and written consent, we interviewed participants about background factors and about illness and mortality in their home since March 25. Two drops of blood obtained by finger prick were applied to a point-of-care antibody test (OnSite COVID-19 IgG/IgM Rapid Test, CTK Biotech).

The study was approved by the Guinean National Ethics Committee (Ref 116/CNES/INASA/2020).

## Results

A total of 146 staff/associated were present to be tested during the survey, 6 declined participation. Of 140 tested, 25 (18%) were IgG-positive. One person was positive for IgM and had a slight fever. The person was referred for PCR testing, which was negative; all symptoms waned after a day.

The average age of IgG-positives tended to be higher than among IgG-negatives (mean 46 years (range 26-70) vs 41 years (range 19-63), p=0.05). There tended to be more infected females than males (24% vs 13%, p=0.10) (Table 1). The ethnicities that traditionally populated the study area had a higher risk of being IgG-positive than other ethnicities. All participants reported using masks.

**Table 1.** Characteristics of individuals testing IgG positive or IgG negative for SARS-CoV2 in Guinea-Bissau, Nov 2020

|  |  | **IgG positive**<br>**(% of group)**<br>**N=25** | **IgG negative**<br>**N=115** | **Relative risk**<br>**(95% CI)** | **P-value** | **Multivariable**<br>**model\*\*\*\*** |
| --- | --- | --- | --- | --- | --- | --- |
| **Mean age in years**<br>**(range)** |  | 46 (26-70) | 41 (19-63) | 1.03 (1.00-1.07) | 0.05 | 1.02 (0.98-1.05) |
| **Sex** | Male | 10 (13%) | 67 | Ref | 0.10 | Ref |
|  | Female | 15 (24%) | 48 | 1.83 (0.88-3.81) |  | 1.21 (0.57-2.60) |
| **Ethnicity\*\*** | Pepel/Manjaco/Mancanha | 20 (27%) | 54 | 3.57 (1.41-9.00) | 0.003 | 3.19 (1.23-8.27) |
|  | Other | 5 (8%) | 61 | Ref |  | Ref |
| **Type of work** | Field assistants | 2 (8%) | 24 | Ref | 0.05 |  |
|  | Office staff | 6 (17%) | 30 | 2.17 (0.47-9.95) |  |  |
|  | Doctors/nurses/midwives | 10 (24%) | 31 | 3.17 (0.75-13.4) |  |  |
|  | Lab technicians | 5 (42%) | 7 | 5.41 (1.21-24) |  |  |
|  | Other staff | 2 (8%) | 23 | 1.04 (0.16-6.87) |  |  |
| **Health care worker** | Yes | 15 (28%) | 38 | 2.46 (1.19-5.09) | 0.01 | 2.22 (1.06-4.67) |
|  | No | 10 (11%) | 77 | Ref |  | Ref |
| **Area of residence\*\*\*** | Bandim/Belém/Mindera/Cuntum | 5 (9%) | 50 | Ref | 0.02 | Ref |
|  | Praça/Antula | 4 (15%) | 23 | 1.62 (0.47-5.61) |  | 1.63 (0.50-5.66) |
|  | Missira/Militar/Aeroporto | 5 (18%) | 23 | 1.96 (0.62-6.25) |  | 2.23 (0.80-6.19) |
|  | Bor/Quelélé/Enterramento | 11 (37%) | 19 | 4.03 (1.54-10.6) |  | 3.17 (1.22-8.22) |
| **Median number of people in household** |  | 5 (1-9) | 5 (1-21) |  | 0.77 |  |
| **Median number of people in house** |  | 10 (3-26) | 12 (1-30) |  | 0.43 |  |
| **Ill during the pandemic** | Yes | 12 (21%) | 45 | 1.34 (0.66-2.74) | 0.42 |  |
|  | No | 13 (16%) | 70 | Ref |  |  |
| **Among the ill** |  | **N=12** | **N=45** |  |  |  |
| - **Loss of taste/smell** | Yes | 7 (28%) | 18 | 1.79 (0.64-5.02) | 0.27 |  |
|  | No | 5 (16%) | 27 | Ref |  |  |
| - **Fever** | Yes | 8 (23%) | 27 | 1.26 (0.42-3.72) | 0.68 |  |
|  | No | 4 (18%) | 18 | Ref |  |  |
| - **Cough** | Yes | 3 (16%) | 16 | 0.67 (0.20-2.20) | 0.61 |  |
|  | No | 9 (24%) | 29 | Ref |  |  |
| - **Runny nose** | Yes | 8 (17%) | 40 | 0.38 (0.14-0.99) | 0.05 |  |
|  | No | 4 (44%) | 5 | Ref |  |  |
| - **Difficulties breathing** | Yes | 2 (33%) | 4 | 1.70 (0.48-6.06) | 0.41 |  |
|  | No | 10 (20%) | 41 | Ref |  |  |
| - **Fatigue** | Yes | 4 (29%) | 16 | 0.90 (0.31-2.65) | 0.85 |  |
|  | No | 8 (20%) | 29 | Ref |  |  |

\*By ranksum test (number of people in household/house) or Poisson test with robust variance estimation (rest)
\*\* Grouped into traditional ethnicities in the study area, with related languages and social structures, vs. others
\*\*\* Grouped by geographical vicinity: the Bandim Health Project study area (Bandim/Belém/Cuntum); areas closer to city (Praça/Antula); areas further from city on the northern side (Missira/Militar/Aeroporto); areas further out of the city on the southern side (Bor/Quelélé/Enterramento)
\*\*\*\*Retaining age, sex, and variables that were significant in univariate analysis

The highest proportion of IgG-positives (42%) was found among laboratory staff, followed by frontline health care workers (HCWs) (doctors, nurses or midwifes) (24%) and office personnel (17%). In the combined group of frontline HCWs and laboratory technicians, 28% tested positive (p for same risk as others=0.01). The lowest proportions were among field assistants (8%) and other staff (mechanics, guards, cleaners) (9%).

The area of residence was associated with the risk of being IgG-positive, the proportion varying from 9% to 37% (p=0.02 for same risk across areas).

In a multivariable analysis retaining age and sex and the three variables (ethnicity, type of work and area of residence) that were significant in univariate analysis, all three variables remained independently associated with the risk of being IgG-positive.

In urban Guinea-Bissau, most people live in multi-family houses. There was no association between being IgG-positive and the number of household inhabitants or the total number of people in the multi-family house (Table 1). Five people reported that a person in their house had died. Five deaths during 8 pandemic months among 140 people, who live in houses with a mean of 12 people/house corresponds roughly to a crude yearly death rate of 4.5/1000 people. No death was attributed to COVID-19.

The risk of being IgG-positive did not correlate with self-reported illness (Table 1). Among the IgG-positives, 12 (48%) reported having been ill since the onset of the pandemic, vs. 45 (40%) of IgG negative (p=0.41). Of the 57 persons reporting being ill, 25 reported loss of smell or taste: 7 of these were IgG-positive (58% of IgG-positives), while 18 were IgG-negative (40% of IgG-negatives)(p=0.27). One person was hospitalized during the pandemic; this person was not PCR-positive, and tested IgG negative here.

Interestingly, 25 (18%) participants had been PCR-tested between March and September; 7 were reported to be positive. Among these 7, 6 reported being ill during the pandemic, all reported loss of taste/smell and runny nose, approx. half reported fever and/or cough; only one reported difficulty breathing and one reported fatigue. Four of the 7 PCR-positives now tested IgG-negative.

## Discussion

COVID-19 infections may no longer be widely transmitted in Bissau, with the apparent decline coinciding with the start of the rainy season in June. In this serosurvey, 18% had IgG antibody.

Only 3 of 7 PCR-positive also tested IgG-positive. Given that the pandemic might have peaked in weeks 17-23 (Figure), most may have been infected more than 5 months before our survey. It has previously been shown that negative SARS-CoV-2 serology does not exclude previous infection[2]. Point-of-care rapid tests are not as precise or sensitive as laboratory antibody tests. The CTK test used for the present study was, however, among the best in a comparison of nine SARS-CoV-2 immunoassays, with a specificity of 90% and a specificity of 100%[3].

**Figure.**
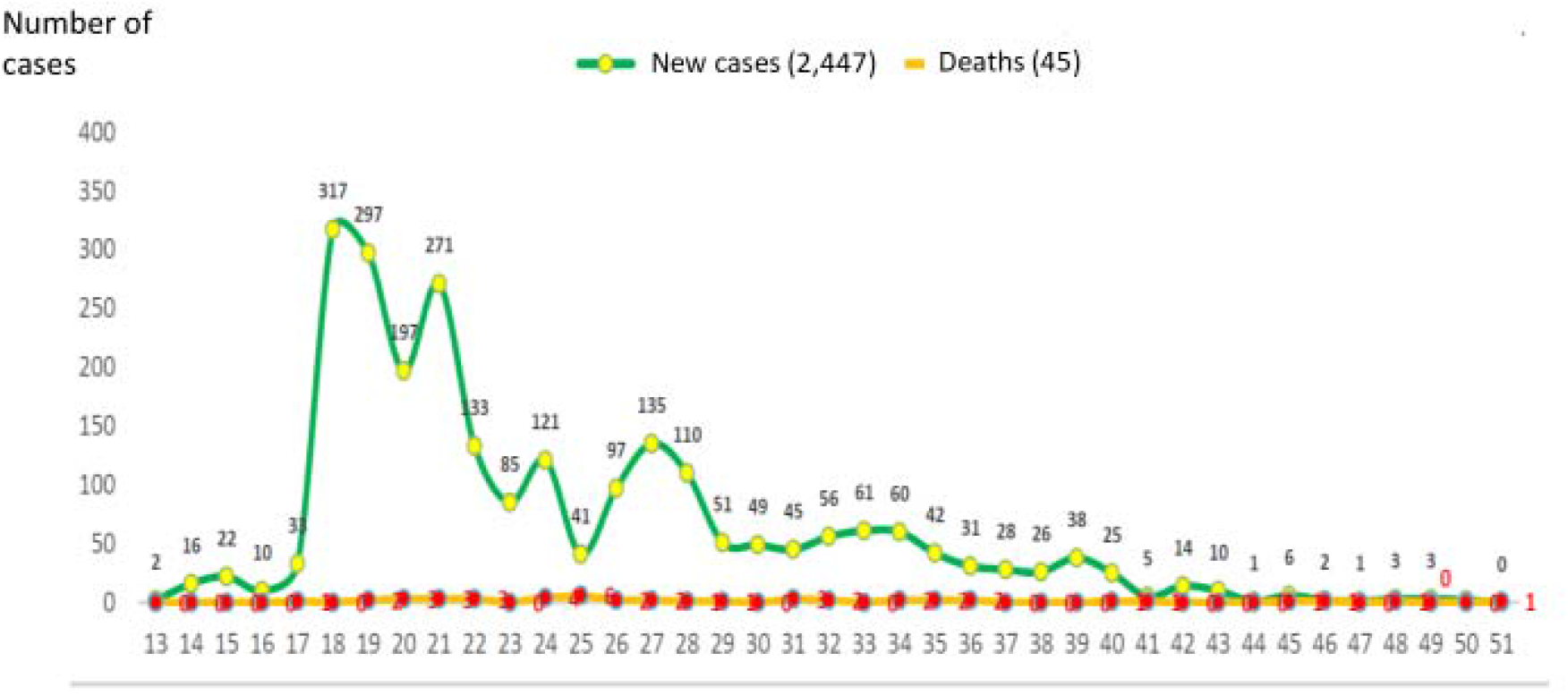
Epidemic curve of registered new cases and deaths from COVID-19 during weeks of 2020 in Guinea-Bissau[11].

The point-of-care test is not a quantitative test, but we noted that many positives exhibited a quite weak, lighter colored IgG band than we have seen for people recently infected in Denmark. Only 12 of the 25 IgG-positive individuals reported being ill during the pandemic. Loss of smell/taste is a common symptom of COVID-19 and was reported by all who had been ill and had a positive PCR test but was also reported by many IgG-negative individuals. Hence, the true prevalence of past infection might be underestimated by IgG-positivity. On the other hand, HCWs were overrepresented in our study population which could overestimate the seroprevalence compared to the general population.

To our knowledge, seven other African SARS-CoV2 serosurveys have been conducted, in Togo[4] (1.6% positive), Ethiopia[5] (3%), Kenya[6] (4.3%), Malawi[7] (12%), South Africa[8] (40%), and Nigeria[9, 10] (25% and 45%)(Supplementary table 1). A common feature seems to be that HCWs had the highest risk. Numbers were small but laboratory workers had the highest risk of all and may be a subgroup that deserves special attention as they collect and process patient samples, but perhaps without the same level of protection as frontline HCWs. Our data suggest that persons working outside may have a lower risk.

The highest risk was seen outside the Bandim Health Project’s study area. This may suggest that infection was more present in some areas and that people to a large extent were infected at home. However, there was no association with the number of people in the household or the house, as would be anticipated if infection at home was prevalent. Alternatively, since the prevalence was highest in the most distant suburbs, shared transport, which mainly consists of crowded minibuses, could be a risk factor.

In conclusion, our survey found a high prevalence of IgG-positive individuals in an urban African setting. COVID-19 was certainly here. The official numbers grossly underestimate the true number of COVID-19 cases. More than half of the IgG-positives had not been ill. Studies are ongoing to assess the overall mortality impact of the pandemic. Despite low official numbers, the toll might have been high, and undetected among the elderly. However, the overall yearly mortality rate of 4.5/1000 people in this study is lower than the official Guinean 2018 yearly mortality rate of 9.6/1000 people[1]. As for now, some degree of herd immunity seems to be present.

## What is already known on this subject

- Many African countries have experienced far fewer COVID-19 cases than countries in Europe, Asia or the Americas.
- By the end of 2020, Guinea-Bissau had <2500 PCR-confirmed cases corresponding to 0.1% of the national population.

## What this study adds

- Among 140 field station staff members, the proportion being SARS-CoV2 IgG positive was 18%.
- Less than half of the IgG-positive individuals reported being ill during the pandemic.
- The official number of PCR-confirmed COVID-19 cases grossly underestimates the prevalence during the pandemic.

## Data Availability

Reg. data sharing, please contact the corresponding author.

## Author Contributions

All authors had had full access to all the data in the study. Benn takes responsibility for the integrity of the data and the accuracy of the data analysis.

## Concept and design

Benn, Cabral, Martins, Schaltz-Buchholzer, Aaby. Acquisition of data: Benn, Salinha, Fernandes.

## Analysis, or interpretation of data

All authors.

## Drafting of the manuscript

Benn, Nielsen, Fisker, Schaltz-Buchholzer, Jørgensen, Aaby.

## Critical revision of the manuscript for important intellectual content

All authors.

## Statistical analysis

Benn.

## Obtained funding

NA

## Administrative, technical, or material support

Cabral, Jørgensen.

## Conflict of Interest Disclosures

No disclosures were reported.

## Funding/Support

EDCTP funded a study of BCG vaccine to health care workers including part of the staff involved in the present survey (RIA2020EF-3049). Statens Serum Institut, Denmark, donated the test kits used while other expenses were covered by in-kind funding. The funders had no role in the design and conduct of the study; collection, management, analysis, and interpretation of the data; preparation, review, or approval of the manuscript; and decision to submit the manuscript for publication.

## Disclaimer

The content of this article is solely the responsibility of the authors and does not necessarily represent the official views of any funder or sponsor.

**Supplementary table 1.** Overview of serosurveys in Sub Saharan Africa published per end of 2020 – sorted chronologically.

|  | Period | Population | Assessment technique | % IgG positivity |  |
| --- | --- | --- | --- | --- | --- |
| Togo[4] | 23 April – 8 May 2020 | High risk population (health care workers, transport sector etc) | Lungene Rapid Test | 1.2% |  |
| Ethiopia[5] | May 2020 | Asymptomatic individuals at a laboratory | Abbott IgG test on ARCHITECT platform | 3% |  |
| Kenya[6] | 30 April – 16 June 2020 | Samples from blood donors | Spike ELISA | 4.3% weighted/adjusted (5.6% crude) |  |
| Malawi[7] | 22 May - 19 June 2020 | Health care workers | ELISA | 12% |  |
| Niger State, Nigeria[10] | 26 June - 30 June 2020 | Health care workers | Colloidal gold rapid test kit | 37% | Total:25% |
|  |  | Non-health-care workers |  | 19% |  |
| South Africa[8] | 15 July - 7 August 2020 | People living with HIV | Elecsys anti-SARS-CoV-2 assay (Roche, Geneva, Switzerland) | 42% | Total: 40% |
|  |  | Pregnant women |  | 38% |  |
| Nigeria[9] | Not reported | Health care workers | ELISA | 45% |  |
| Guinea-Bissau (present study) | 9 - 24 November 2020 | Health care workers and field station staff | CTK POC rapid test | 18% |  |

## Notes

### Competing Interest Statement

The authors have declared no competing interest.

### Author Declarations

The study was approved by the Guinean National Ethics Committee (Ref 116/CNES/INASA/2020).

